# Temporal Changes in Youth Subjective Well-Being in Nigeria, 2016–2021

**DOI:** 10.64898/2026.01.20.26344435

**Authors:** Kamaldeen Sunkanmi Abdulraheem

**Affiliations:** Department of Community Health and Primary Health, Lagos University Teaching Hospital, Idi-Araba, Surulere, Lagos, Nigeria.

**Author notes:** Corresponding Author: Kamaldeen Sunkanmi Abdulraheem Department of Community Health and Primary Health, Lagos University Teaching Hospital, Idi-Araba, Lagos State, Nigeria.

**Keywords:** Nigeria, Youth, Subjective well-being, Happiness, Life satisfaction, Subnational inequality, Micro–macro linkage, Fragile settings

## Abstract

This study examines the impact of subnational structural determinants on the subjective well-being of Nigerian youth (aged 15–24) between 2016 and 2021. Individual-level data from IPUMS-harmonised Multiple Indicator Cluster Surveys (MICS) were linked to state-year contextual indicators from the Global Data Lab, including the Global Vulnerability Index (GVI), a corruption-based institutional quality measure, and the Gini coefficient of income inequality. This micro–macro linkage situates youth well-being within social, institutional, and economic environments. Survey-weighted regression models accounting for the complex MICS sampling design reveal a significant decline in affective well-being over time, with the 2021 period effect remaining robust after adjustment for institutional quality, fragility, and inequality. Female gender and higher education emerged as protective factors, while formerly or never-married status increased vulnerability. By integrating individual survey data with spatially indexed structural indicators, the study demonstrates the analytical value of micro–macro linkages for understanding how subnational contexts shape youth well-being in fragile and unequal settings.

## Introduction

Subjective well-being (SWB)—encompassing both affective components (happiness, positive and negative emotions) and evaluative components (life satisfaction and cognitive judgments about one’s life)—is a widely recognised indicator of quality of life and psychological health.^1–3^ Beyond individual mental states, SWB captures how people experience and appraise their lives within broader social, economic, and institutional contexts. While SWB has been extensively studied in high-income countries, it remains comparatively under-researched in low- and middle-income countries (LMICs), particularly in sub-Saharan Africa (SSA), where persistent structural challenges—including inequality, institutional fragility, and exposure to recurrent shocks—may profoundly shape how young people perceive their lives.^4^ Evidence from SSA further suggests that social and economic inequalities are closely associated with variations in subjective well-being, underscoring the importance of contextual and structural conditions in shaping life satisfaction and happiness.^5^

In Nigeria, youth aged 15–24 constitute a critical demographic group, accounting for a substantial share of the population, with approximately 58% under the age of 30 and a median age of 18.1 years.^6^ he well-being of this cohort is central to the country’s prospects for harnessing a demographic dividend, yet it is threatened by systemic constraints, including limited access to quality education and healthcare, economic volatility, unemployment, and pervasive governance challenges.^7^ Existing Nigerian research on SWB has largely concentrated on individual-level or psychosocial correlates—such as material conditions, psychological well-being, or family relationships—with comparatively limited attention to how broader state-level structural environments shape youth well-being.^8,9^ As a result, important dimensions of inequality in lived experience across Nigeria’s highly heterogeneous states remain insufficiently understood.

This gap is particularly consequential given the scarcity and fragmentation of population-level mental health data in Nigeria, especially within nationally representative surveys.^10^ In this context, SWB represents one of the most feasible and policy-relevant population-level proxies for mental health–related outcomes, capturing individuals’ cognitive and affective evaluations of their life circumstance.^11^ Although SWB does not replace clinical diagnoses or fully capture psychiatric morbidity, it is sensitive to social, economic, and institutional conditions and has been shown to correlate strongly with broader indicators of psychological well-being.^12^ Large-scale household surveys such as the Multiple Indicator Cluster Surveys (MICS) routinely collect SWB-related measures, providing a rare opportunity to examine population well-being across subnational units and over time in LMIC settings.

Between 2016 and 2021, Nigeria experienced a series of overlapping macroeconomic and social disruptions, including economic recession, oil price volatility, rising insecurity in several regions, and a major global public health shock.^13^ These disruptions unfolded unevenly across states, amplifying pre-existing differences in governance capacity, structural vulnerability, and inequality. Yet, despite growing global evidence that macro-level factors—such as income inequality and institutional quality—exhibit context-dependent and sometimes counterintuitive relationships with SWB, Nigeria lacks integrated empirical analyses linking such structural indicators to nationally representative individual-level data.^14^ Understanding how these contextual factors shape youth well-being is especially important in fragile and unequal settings, where structural conditions may either buffer or exacerbate the psychological consequences of social change.

Against this backdrop, large-scale household surveys such as the MICS offer a unique opportunity to examine youth subjective well-being across temporal and subnational contexts in Nigeria. By linking individual-level SWB data with state-level indicators of institutional quality, structural vulnerability, and inequality, it becomes possible to move beyond purely individual explanations and assess how place-based conditions shape well-being trajectories.

### Objective

The objective of this study was to link nationally representative IPUMS-harmonised MICS data with subnational structural indicators to examine temporal changes in youth subjective well-being in Nigeria between 2016 and 2021, and to assess how state-level institutional quality, structural vulnerability, and inequality are associated with affective and evaluative dimensions of well-being during this period.

## Methodology

### Data Sources and Sampling Frame

This study employs a cross-sectional, comparative design using two waves of the Multiple Indicator Cluster Surveys (MICS) for Nigeria, accessed via IPUMS: MICS5 (2016–2017) and MICS6 (2021). These nationally representative household surveys, developed by UNICEF, monitor the situation of children, women, and (increasingly) men.^9,10^

Both waves used a two-stage stratified cluster design, with states (nested within six geopolitical zones) as main strata. Enumeration areas (EAs; primary sampling units) were selected systematically with probability proportional to size from census-based master samples. Households were then systematically sampled after listing (16 per EA in 2016–2017; 20 per EA in 2021). The 2021 wave included a larger supplemental sample (337 clusters/6,740 households) for the integrated National Immunization Coverage Survey (NICS), with partial questionnaires administered therein. Insecurity limited access in Northeast states (e.g., 101 EAs in 2016–2017; 128 in 2021, with Borno restricted to accessible local government areas covering 29% population). Samples are not self-weighting; design weights adjust for non-response.^15–17^

The analytic subsample comprises youth aged 15–24 completing Individual Questionnaires for Women or Men (total unweighted N=41,175). All descriptive statistics and regression models use survey weights to produce nationally representative estimates.

## Study Variables

### 2.2 Measurement of Subjective Well-Being (SWB)

The primary outcome was affective well-being (overall happiness), measured consistently across waves. Evaluative well-being (life satisfaction) was a secondary outcome, analyzed separately due to measurement change. In the 2016–2017 wave, respondents were asked: “How satisfied are you with your life, overall?” using a 5-point verbal scale (1 = very satisfied to 5 = very unsatisfied).^16^ In the 2021 wave, the Cantril Self-Anchoring Ladder was introduced: respondents indicated the step (0 = worst possible life to 10 = best possible life) on which they currently stand.^15^ The overall estimation of happiness (“Taking all things together, would you say you are very happy, somewhat happy, neither happy nor unhappy, somewhat unhappy, or very unhappy?”) remained identical in wording and response format across both waves, allowing reliable examination of temporal trends in affective well-being.^15,16^ In contrast, the change in the evaluative life satisfaction question precludes direct comparison of trends between waves. However, the consistent happiness measure enables robust assessment of the impact of state-level contextual factors (Global Vulnerability Index, Cleanliness Index, and Gini coefficient) and their interaction with time on youth well-being.

### 2.3 Individual-Level Predictors (Covariates)

To minimize omitted variable bias and isolate the effects of state-level structural conditions, a comprehensive set of socio-demographic and psychological covariates was harmonized across both survey waves. These included age (continuous, mean ≈18.9 years), gender (binary: Male [reference], Female), education (three-level factor: Primary or less [reference], Secondary, Tertiary; unknown cases retained as a separate category to preserve sample size), residence (binary: Urban [reference], Rural), marital status (three-level factor: Married/In Union [reference], Formerly Married, Never Married), and ethnicity (household head ethnicity: Hausa [reference], Igbo, Yoruba, Other) as a proxy for cultural and regional confounding. Additionally, two items capturing perceived well-being trends—comparison to the previous year (“Improved” [reference], “More or less the same,” “Worsened”) and expectations for the next year (“Better” [reference], “More or less the same,” “Worse”)—were included to account for individual psychological trajectories and potential mediation of period effects.^9,10^

### 2.4 State-Level Contextual Indicators (GDL Data)

To situate individual-level responses within broader structural contexts, survey records from IPUMS MICS were linked to subnational macro-indicators from the Global Data Lab (GDL) Subnational Human Development Database.^18^ The linkage was performed via a many-to-one merge at the state level (N=37 units, including the Federal Capital Territory) using geographic region codes and survey year as keys. This approach allowed alignment of individual observations with the corresponding state-year values of the following indicators:

- **Institutional Context** — Corruption Index (fullsci): a composite measure of institutional quality, reflecting the absence of corruption and bureaucratic effectiveness. Higher values indicate better institutional performance.
- **Fragility Context** — Global Vulnerability Index (GVI): a multidimensional index capturing exposure to structural risks, including environmental, economic, and social shocks. Higher values denote greater vulnerability.
- **Inequality Context** — Gini coefficient: a standard measure of income inequality within states, ranging from 0 (perfect equality) to 1 (perfect inequality).

Due to high collinearity between GVI and the Gini coefficient (Pearson r ≈ 0.87), these indicators were tested in separate models, with GVI retained as the primary fragility measure in final specifications. All GDL indicators are publicly available, annually updated, and designed for subnational comparative analysis across countries.^18^

This micro–macro linkage enables the examination of how place-based structural conditions moderate the relationship between national period effects and individual youth well-being.

### 2.5 Data Preparation and Merging

Prior to data analysis, the raw MICS data from IPUMS were processed in Stata to merge gender-specific modules for men and women. Specifically, individual questionnaires for men (containing variables such as edlevelmn for education and agemn for age) and women (edlevelwm and agewm) were combined using a unique household and person ID key (e.g., using Stata’s merge 1:1 hh_id person_id using "women_data.dta" command). This created a unified dataset with coalesced variables (e.g., education = edlevelmn if male, edlevelwm if female; age = agemn if male, agewm if female). Missing or NIU (not in universe) values were retained during merging to preserve sample integrity. The merged Stata dataset was then exported as a .dta file and imported into R for further cleaning, harmonisation, and subsetting to youth aged 15–24 (using haven package).

### 2.6 Statistical Analysis and Survey Weights

All analyses were conducted using R version 4.4.1 using the survey and gtsummary packages. To properly account for the complex multi-stage stratified cluster design of the MICS surveys, a survey design object was constructed using the svydesign function. The design incorporated primary sampling units (PSUs; census enumeration areas) as clustering IDs, states as stratification variables, and normalized survey weights. Raw weights were rescaled such that their sum equaled the actual analytic sample size (n = 41,175), a standard adjustment that prevents artificial inflation of precision while fully preserving design-based representativeness and variance structure.^19^ Nesting was enabled (nest = TRUE) to correctly handle the reuse of PSU identifiers across state-level strata, ensuring accurate standard errors for all estimates.

### 2.7 Modeling Strategy

All models were estimated using design-based generalized linear models (svyglm, Gaussian family) for the continuous affective happiness outcome (reversed 1–5 scale; evaluative life satisfaction was analyzed separately by year).

- **Phase 1 – Structural Models**: Baseline models of the form y = β₀ + β₁(Year) + β₂(Demographics) + β₃(MacroIndicator) + ε were estimated, where macro-indicators (Cleanliness Index, GVI, Gini) were tested in separate specifications to avoid multicollinearity (e.g., GVI–Gini correlation r ≈ 0.87). These models quantified the raw 2016–2021 decline and the direct contribution of each structural context, net of individual demographics.
- **Phase 2 – Resilience Models**: The structural models were extended by adding psychological and social buffers (perceived past/future well-being trends, marital status, ethnicity) to assess the extent to which individual-level resilience factors mediate the period effect and attenuate the association between state vulnerability and life satisfaction.
- **Phase 3 – Interaction Models**: To test the hypothesis of an “erosion of resilience,” a key interaction term was introduced:

y = β₀ + β₁(Year) + β₂(GVI) + β₃(Year × GVI) + γ(Demographics + Resilience) + ε This specification examined whether the negative effect of state-level vulnerability (GVI) on youth subjective well-being intensified in 2021 compared with 2016.

As a robustness check, random-intercept linear mixed-effects models were estimated using the lme4 package, treating states as random effects. The intraclass correlation coefficient (ICC = σ²_state / (σ²_state + σ²_residual) was calculated to quantify the proportion of variance in life satisfaction attributable to between-state differences versus individual variation. Results were substantively consistent with the primary design-based models, supporting the use of survey-weighted fixed-effects specifications as the main approach. P-values and 95% confidence intervals are design-based to reflect the complex sampling structure. Statistical significance was defined as p < 0.05 (two-sided).

### 2.8 Ethical Considerations

This research uses secondary, de-identified data from two rounds of the Nigeria Multiple Indicator Cluster Surveys (MICS), harmonized and distributed by IPUMS MICS. The original MICS protocols, implemented by the National Bureau of Statistics (NBS) with UNICEF technical support, included ethical safeguards such as verbal informed consent from respondents, advance adult consent and child assent for youth aged 15–17, assurances of voluntary participation, confidentiality, anonymity, and the right to refuse questions or terminate interviews at any time. A protection protocol addressed potential risks throughout the survey lifecycle. No primary data collection occurred in this study, and all records were fully anonymized with no identifiable information. Therefore, no further ethical review or approval was required

## Results

Youth happiness (reversed scored) declined from a mean of 4.5 (SD 0.8) in 2016 to 4.1 (SD 1.0) in 2021 (p < 0.001). The proportion reporting “very happy” fell from 61% to 41%, while neutral and unhappy categories increased (p < 0.001). Evaluative life satisfaction showed a mean verbal score of 1.6 (SD 0.9) in 2016 (1–5 scale) and a mean Cantril Ladder score of 6.2 (SD 4.6) in 2021 (0–10 scale; direct comparison not possible due to different instruments). The sample was 68% female, with a mean age of 18.9 years (SD 2.8). Education levels rose slightly (tertiary from 7.7% to 10%), never-married youth increased from 69% to 79% (p < 0.001), and rural residence decreased from 64% to 56% (p = 0.002). Perceptions of past well-being worsened (improved from 73% to 63%, p < 0.001), as did future expectations (better from 91% to 88%, p < 0.001). Ethnic composition shifted, with fewer Hausa-headed households in 2021 (29% vs 48%, p < 0.001). Frequency of TV watching remained stable, with “not at all” being the most common response (46% overall)

In 2016, higher verbal life satisfaction was associated with female gender, older age, formerly/never married status, and positive past well-being perceptions, while tertiary education was negatively associated. In 2021, female gender and tertiary education were strongly positive, whereas formerly married status and worsened past perceptions were strongly negative; future expectations showed consistent associations across years.

Table 3 presents the pooled survey-weighted regression of affective happiness on survey year, state-level fragility, and individual covariates. The survey-weighted regression model (pooled n = 41,160) showed that affective happiness (reversed 1–5 scale, higher = happier) was significantly higher in 2021 than in 2016 after adjustment (β = 0.28, 95% CI 0.10–0.46, p = 0.003), with a positive main effect of state-level fragility (GVI; β = 0.007, p < 0.001). Female gender, secondary and tertiary education were positively associated with happiness, while formerly/never married status, non-Hausa ethnicity, less frequent TV watching, and worsened past well-being perceptions were strongly negative. The model fit was good (AIC = 100,052; residual deviance = 27,262 on 3,757 df).

**Table 1:**
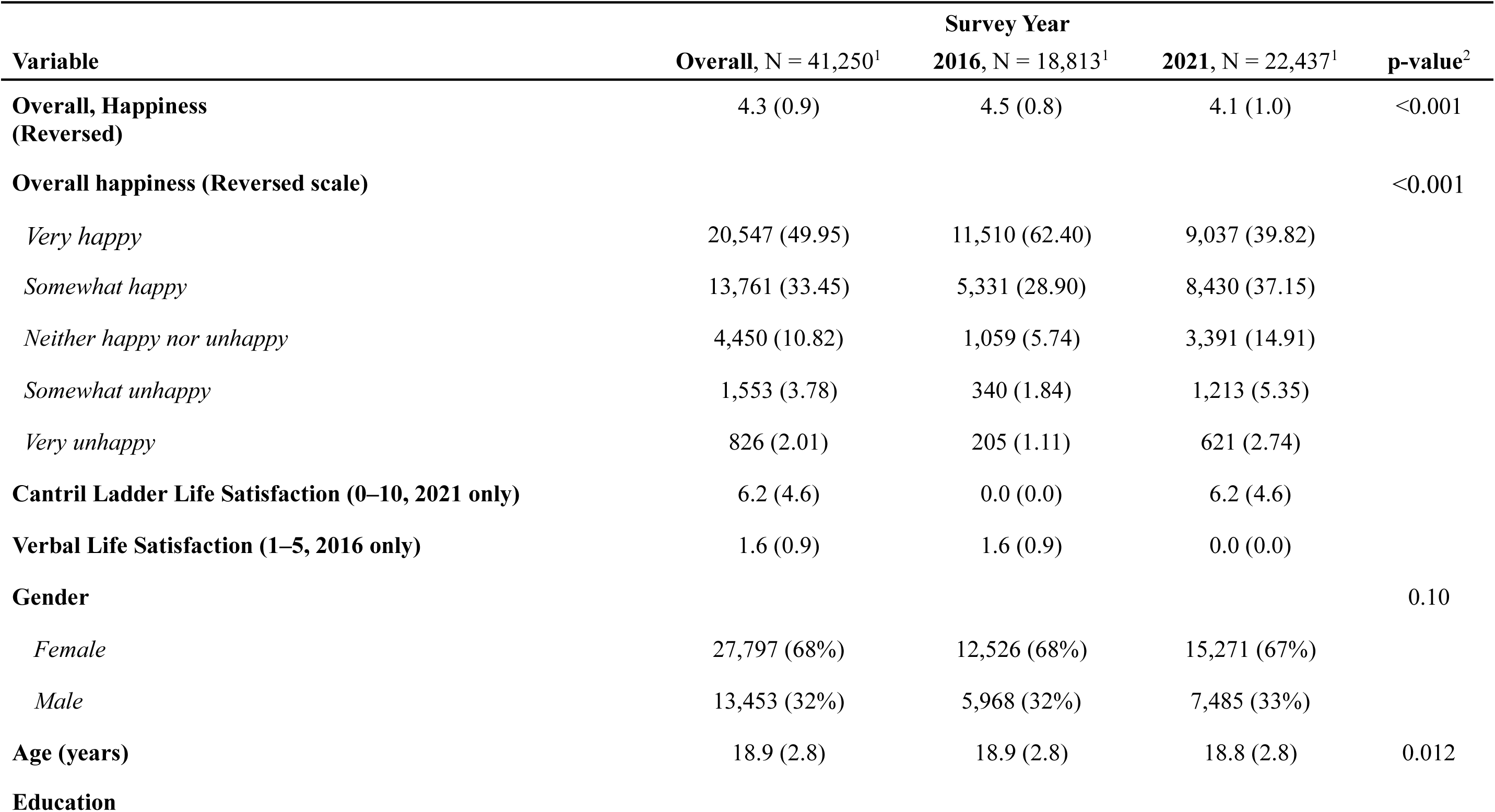
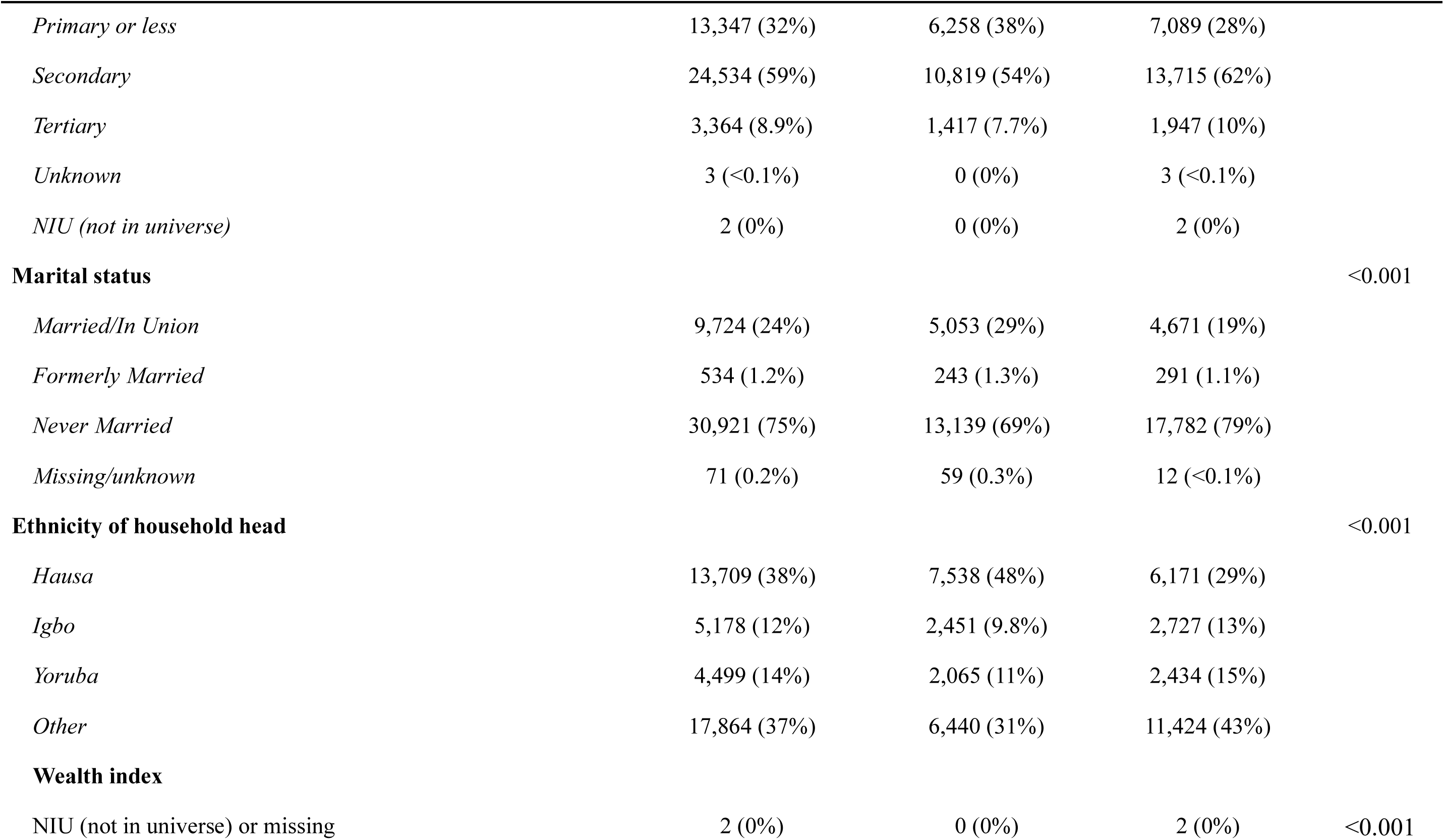
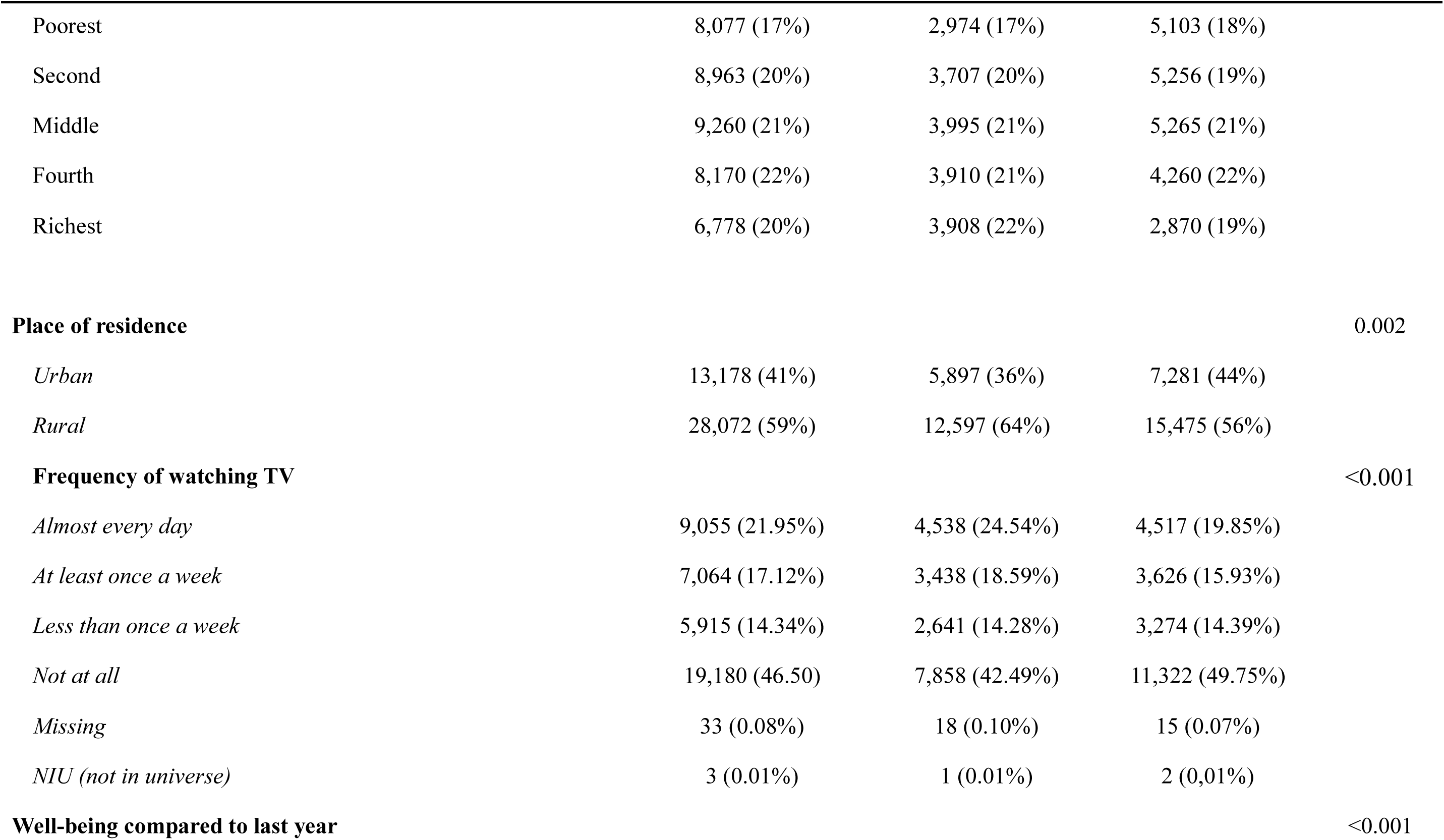
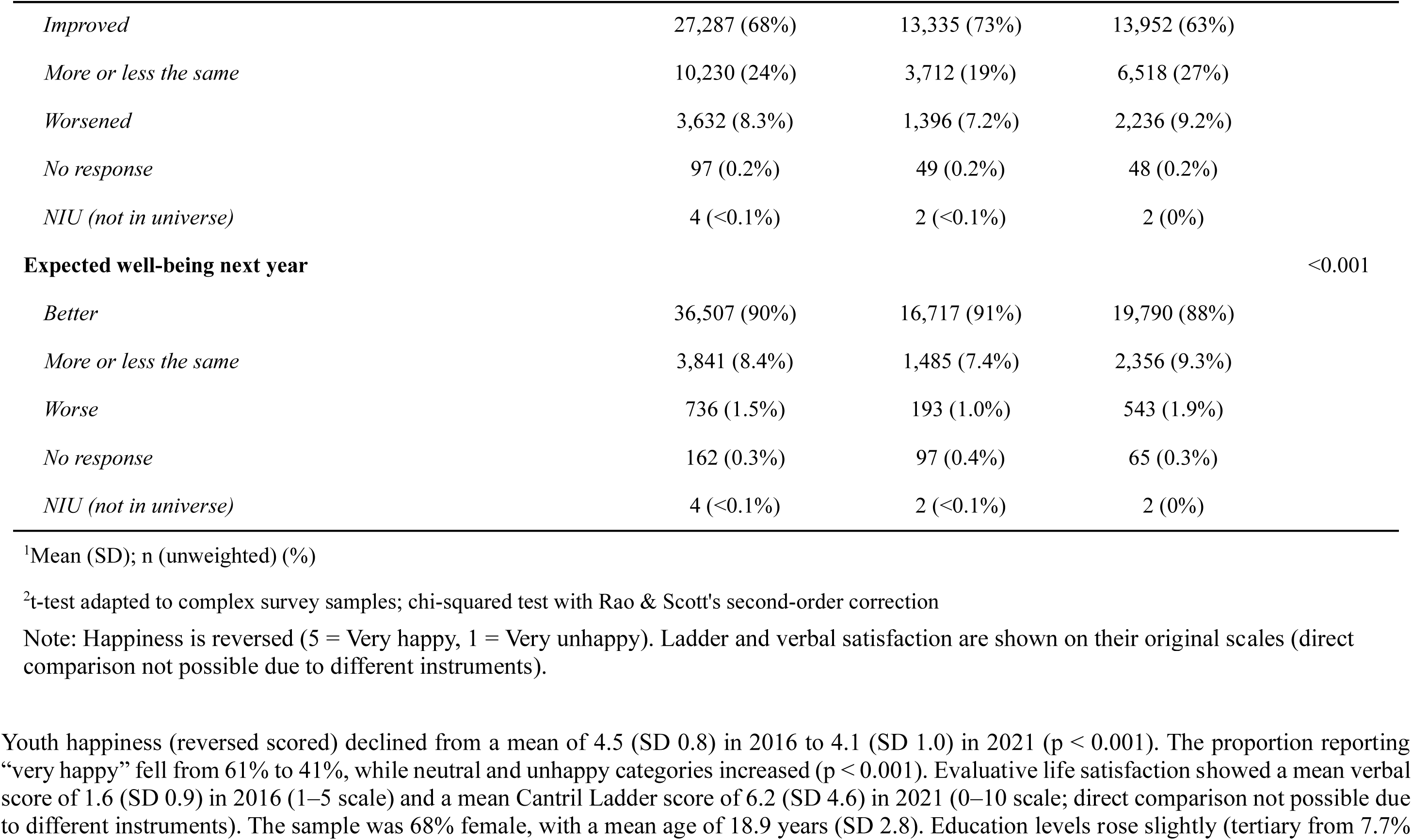
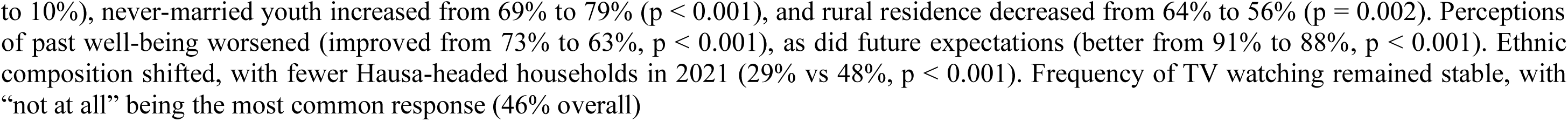
Sociodemographic Covariates and Subjective wellbeing by survey year (2016 and 2021)

**Table 2:** Association between sociodemographic variables and satisfaction with life in Nigeria, 2016 vs 2021

|  | 2016 (Verbal Satisfaction) |  |  | 2021 (Cantril Ladder) |  |  |
| --- | --- | --- | --- | --- | --- | --- |
| Variable | Beta | 95% CI<sup>1</sup> | p-value | Beta | 95% CI<sup>1</sup> | p-value |
| **Gender** |  |  |  |  |  |  |
| Male | — | — |  | — | — |  |
| Female | 0.04 | -0.01, 0.09 | 0.092 | 1.2 | 0.89, 1.5 | <0.001 |
| Age (years) | 0.02 | 0.01, 0.02 | <0.001 | -0.04 | -0.10, 0.02 | 0.2 |
| **Education** |  |  |  |  |  |  |
| Primary or less | — | — |  | — | — |  |
| Secondary | -0.09 | -0.15, -0.03 | 0.002 | 0.26 | -0.05, 0.57 | 0.10 |
| Tertiary | -0.16 | -0.25, -0.08 | <0.001 | 0.50 | 0.07, 0.93 | 0.022 |
| Unknown |  |  |  | -0.69 | -2.9, 1.5 | 0.5 |
| **Marital status** |  |  |  |  |  |  |
| Married/In Union | — | — |  | — | — |  |
| Formerly Married | 0.26 | 0.12, 0.41 | <0.001 | -0.80 | -1.2, -0.40 | <0.001 |
| Never Married | 0.11 | 0.06, 0.15 | <0.001 | 0.03 | -0.27, 0.32 | 0.9 |
| Missing/unknown | 0.11 | -0.15, 0.38 | 0.4 | 0.37 | -0.72, 1.5 | 0.5 |
| **Ethnicity** |  |  |  |  |  |  |
| Hausa | — | — |  | — | — |  |
| Igbo | 0.19 | 0.12, 0.25 | <0.001 | -0.74 | -1.3, -0.20 | 0.007 |
| Yoruba | 0.17 | 0.10, 0.24 | <0.001 | -0.11 | -0.69, 0.47 | 0.7 |
| Other | 0.10 | 0.05, 0.15 | <0.001 | -0.28 | -0.69, 0.13 | 0.2 |
| **Wealth index** |  |  |  |  |  |  |
| Poorest | — | — |  | — | — |  |
| Second | 0.06 | 0.01, 0.12 | 0.025 | -0.23 | -0.64, 0.18 | 0.3 |
| Middle | 0.11 | 0.05, 0.17 | <0.001 | -0.37 | -0.80, 0.07 | 0.10 |
| Fourth | 0.08 | 0.01, 0.14 | 0.017 | -0.29 | -0.75, 0.18 | 0.2 |
| Richest | 0.05 | -0.02, 0.13 | 0.2 | -0.21 | -0.67, 0.25 | 0.4 |
| **Media exposure (television)** |  |  |  |  |  |  |

| Variable | 2016 (Verbal Satisfaction) |  |  | 2021 (Cantril Ladder) |  |  |
| --- | --- | --- | --- | --- | --- | --- |
|  | Beta | 95% CI<sup>1</sup> | p-value | Beta | 95% CI<sup>1</sup> | p-value |
| Almost every day | — | — |  | — | — |  |
| At least once a week | 0.08 | 0.02, 0.13 | **0.004** | -0.15 | -0.72, 0.42 | 0.6 |
| Less than once a week | 0.11 | 0.05, 0.16 | **<0.001** | -0.25 | -0.64, 0.14 | 0.2 |
| Not at all | 0.06 | 0.00, 0.11 | **0.046** | -0.29 | -0.67, 0.10 | 0.15 |
| Missing | 0.30 | -0.08, 0.67 | 0.12 | -1.1 | -2.4, 0.13 | 0.080 |
| **Place of residence** |  |  |  |  |  |  |
| Urban | — | — |  | — | — |  |
| Rural | -0.02 | -0.07, 0.04 | 0.5 | -0.19 | -0.55, 0.18 | 0.3 |
| **Well-being compared to last year** |  |  |  |  |  |  |
| Improved | — | — |  | — | — |  |
| More or less the same | 0.33 | 0.28, 0.37 | **<0.001** | -0.59 | -0.88, -0.30 | **<0.001** |
| Worsened | 0.69 | 0.61, 0.77 | **<0.001** | -1.5 | -1.7, -1.2 | **<0.001** |
| No response | 1.1 | 0.16, 2.0 | **0.022** | 14 | 2.4, 27 | **0.019** |
| **Expected well-being next year** |  |  |  |  |  |  |
| Better | — | — |  | — | — |  |
| More or less the same | 0.12 | 0.06, 0.19 | **<0.001** | -0.34 | -0.59, -0.09 | 0.007 |
| Worse | 0.25 | 0.08, 0.43 | **0.005** | -0.79 | -1.2, -0.35 | **<0.001** |
| No response | 0.28 | -0.02, 0.58 | 0.070 | 14 | 5.3, 23 | **0.002** |

<sup>1</sup>CI = Confidence Interval

**Table 3:** Survey-weighted regression of affective happiness on survey year, state-level fragility (GVI), and individual covariates among Nigerian youth (IPUMS-MICS 2016–2021)

| Characteristic | Beta | 95% CI<sup>1</sup> | p-value |
| --- | --- | --- | --- |
| Year |  |  |  |
| *2016* | — | — |  |
| *2021* | 0.28 | 0.10, 0.46 | **0.003** |
| GVI | 0.01 | 0.00, 0.01 | **<0.001** |
| Gender |  |  |  |
| *Male* | — | — |  |
| *Female* | 0.16 | 0.12, 0.19 | **<0.001** |
| Age (years) | -0.01 | -0.02, -0.01 | **<0.001** |
| Education status |  |  |  |
| *Primary or less* | — | — |  |
| *Secondary* | 0.08 | 0.05, 0.12 | **<0.001** |
| *Tertiary* | 0.11 | 0.04, 0.17 | **0.001** |
| *Unknown* | 0.56 | -0.01, 1.1 | 0.054 |
| Marital status |  |  |  |
| *Married/In Union* | — | — |  |
| *Formerly Married* | -0.41 | -0.54, -0.28 | **<0.001** |
| *Never Married* | -0.08 | -0.12, -0.04 | **<0.001** |
| *Missing/unknown* | 0.11 | -0.08, 0.31 | 0.2 |
| Ethnicity of household head, |  |  |  |
| *Hausa* | — | — |  |
| *Igbo* | -0.06 | -0.13, 0.01 | 0.075 |
| *Yoruba* | -0.05 | -0.12, 0.01 | 0.087 |
| *Other* | -0.04 | -0.08, 0.00 | 0.079 |
| Wealth index |  |  |  |
| *Poorest* | — | — |  |
| *Second* | -0.03 | -0.07, 0.01 | 0.2 |
| *Middle* | -0.01 | -0.06, 0.04 | 0.7 |
| *Fourth* | -0.02 | -0.08, 0.03 | 0.4 |
| *Richest* | 0.01 | -0.05, 0.07 | 0.8 |
| Media exposure (watches television) |  |  |  |
| *Almost every day* | — | — |  |
| *At least once a week* | -0.04 | -0.09, 0.00 | 0.078 |
| *Less than once a week* | -0.12 | -0.17, -0.07 | **<0.001** |
| *Not at all* | -0.01 | -0.06, 0.04 | 0.7 |
| *Missing* | -0.15 | -0.48, 0.18 | 0.4 |
| Place of residence |  |  |  |
| *Urban* | — | — |  |
| *Rural* | 0.03 | -0.02, 0.07 | 0.3 |
| Well-being compared to last year |  |  |  |
| *Improved* | — | — |  |
| *More or less the same* | -0.47 | -0.50, -0.43 | **<0.001** |
| *Worsened* | -0.96 | -1.0, -0.88 | **<0.001** |
| *No response* | -0.92 | -1.3, -0.53 | **<0.001** |
| Expected well-being next year |  |  |  |
| *Better* | — | — |  |
| *More or less the same* | -0.12 | -0.17, -0.08 | **<0.001** |
| *Worse* | -0.43 | -0.60, -0.26 | **<0.001** |
| *No response* | -0.19 | -0.49, 0.10 | 0.2 |
| No. Obs. | 41,160 |  |  |

Model: Survey-weighted generalized linear model (Gaussian family) on pooled 2016–2021 data (n = 41,160 after excluding 90 missing observations). AIC = 100,052; residual deviance = 27,262 on 3,757 df.
<sup>1</sup>CI = Confidence Interval

**Table 4:**
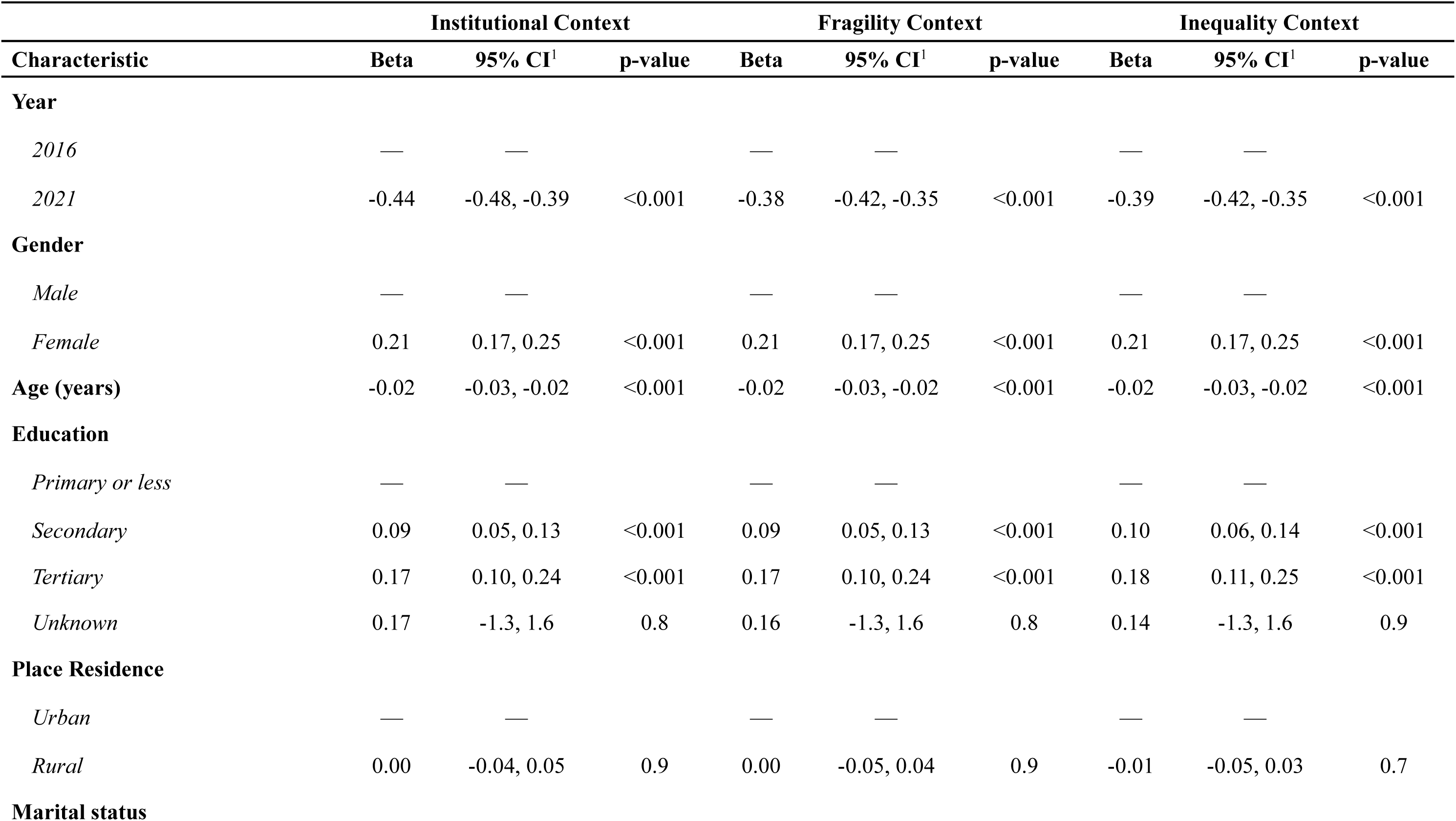
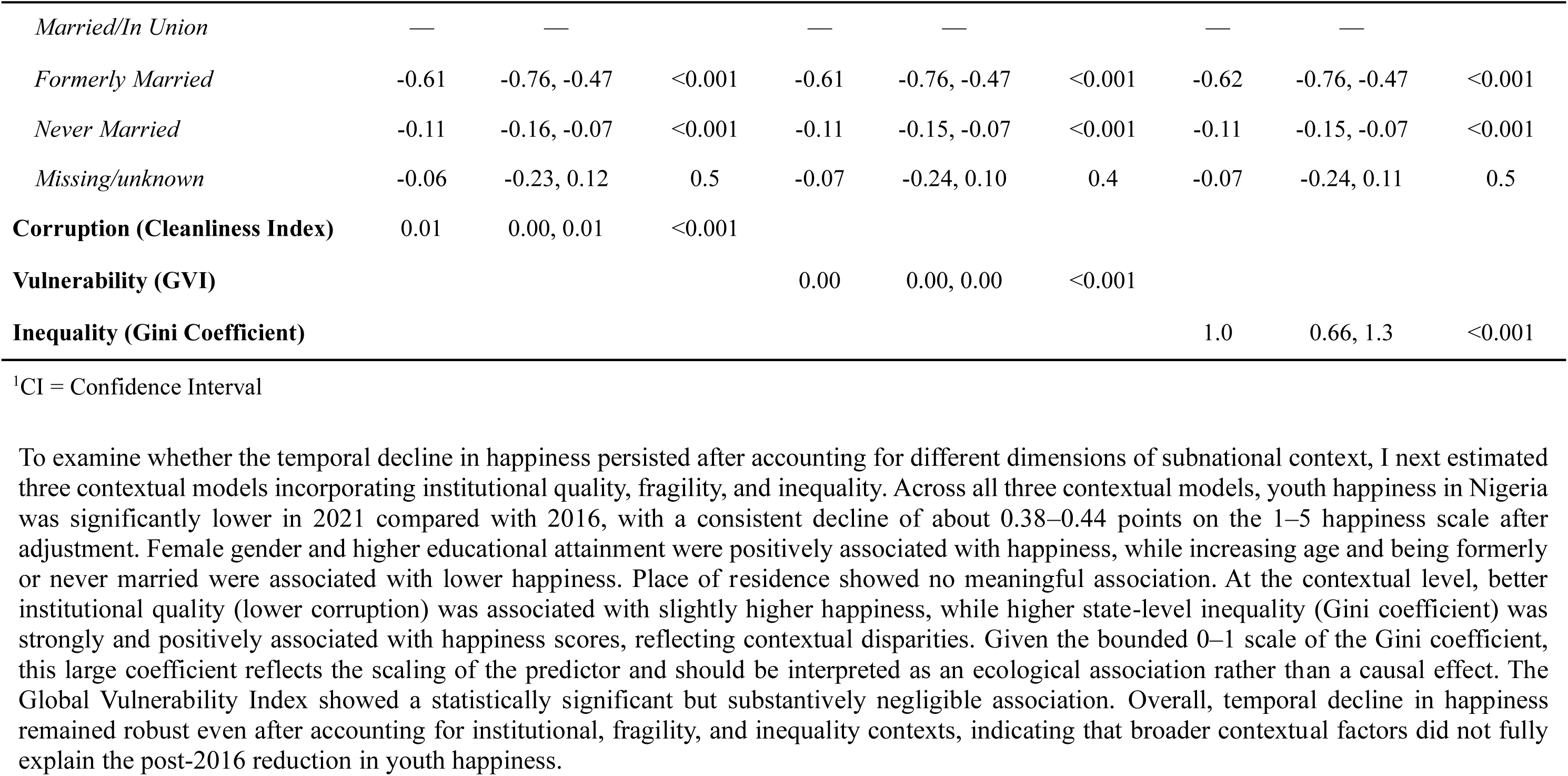
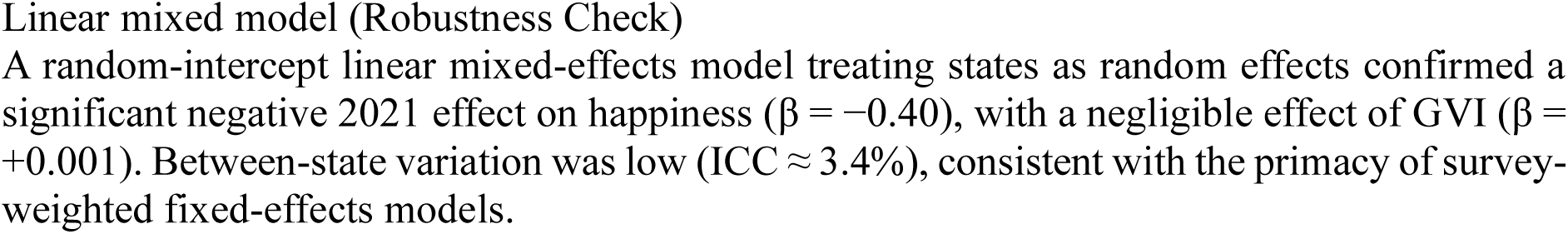
Association between institutional quality, state-level fragility, inequality, and estimation of youth happiness in Nigeria, 2016–2021

To examine whether the temporal decline in happiness persisted after accounting for different dimensions of subnational context, I next estimated three contextual models incorporating institutional quality, fragility, and inequality. Across all three contextual models, youth happiness in Nigeria was significantly lower in 2021 compared with 2016, with a consistent decline of about 0.38–0.44 points on the 1–5 happiness scale after adjustment. Female gender and higher educational attainment were positively associated with happiness, while increasing age and being formerly or never married were associated with lower happiness. Place of residence showed no meaningful association. At the contextual level, better institutional quality (lower corruption) was associated with slightly higher happiness, while higher state-level inequality (Gini coefficient) was strongly and positively associated with happiness scores, reflecting contextual disparities. Given the bounded 0–1 scale of the Gini coefficient, this large coefficient reflects the scaling of the predictor and should be interpreted as an ecological association rather than a causal effect. The Global Vulnerability Index showed a statistically significant but substantively negligible association. Overall, temporal decline in happiness remained robust even after accounting for institutional, fragility, and inequality contexts, indicating that broader contextual factors did not fully explain the post-2016 reduction in youth happiness.

Linear mixed model (Robustness Check) A random-intercept linear mixed-effects model treating states as random effects confirmed a significant negative 2021 effect on happiness (β = −0.40), with a negligible effect of GVI (β = +0.001). Between-state variation was low (ICC ≈ 3.4%), consistent with the primacy of survey-weighted fixed-effects models.

## DISCUSSION

This study examined temporal differences in youth subjective well-being in Nigeria between 2016–2017 and 2021, using affective well-being (overall happiness) as the primary outcome and evaluative well-being (life satisfaction) as a complementary measure. The findings reveal a clear and robust decline in affective happiness over this period, with mean reversed happiness scores and a substantial reduction in the proportion of youth reporting being “very happy”.

This pattern aligns with emerging evidence of worsening mental health among adolescents in SSA, where a meta-analysis reported pooled prevalence rates of 15% for depression and 12% for post-traumatic stress disorder among youth, with higher rates in conflict-affected regions.^20^

Importantly, this decline persisted across multiple models adjusting for state-level institutional quality, fragility, and inequality and sociodemographic factors. While these contextual indicators are theoretically relevant to population mental health, their inclusion did not attenuate the observed temporal decline, suggesting that broader national shocks, cohort-specific stressors, or unmeasured social determinants (including the COVID-19 pandemic and related economic and social disruptions) may be more salient drivers of deteriorating youth affective well-being during this period. Taken together, these findings highlight a concerning downward trend in youth happiness in Nigeria and underscore the utility of subjective well-being measures as proxy indicators for population mental health in settings lacking comprehensive psychiatric surveillance systems.

A particularly salient finding was the consistent negative period effect observed in 2021 (β ≈ −0.38 to −0.44 across contextual models), which emerged most clearly when institutional corruption or quality (Cleanliness Index) or inequality (Gini coefficient) was included in the specification. In contrast, models that included only GVI with sociodemographic covariates produced a counterintuitive positive year effect (β = +0.28) likely reflecting suppression or confounding by omitted contextual variables. This pattern supports the interpretation that the raw descriptive decline is real and not merely an artifact of sample composition change, but also highlights the sensitivity of period estimates to which structural indicators are controlled for.

In contrast, evaluative well-being (overall life satisfaction) could not be directly compared across waves due to the change from a 5-point verbal satisfaction scale in 2016 to the 0–10 Cantril Ladder in 2021. The separate-year models revealed distinct patterns, with happiness and life satisfaction questions correlating moderately in comparable datasets (r ≈ 0.50–0.56) yet capturing related but distinct dimensions.^21,22^ Accordingly, associations were examined separately by survey year. In 2016, higher verbal life satisfaction was associated with older age, formerly or never married status, non-Hausa ethnicity, intermediate wealth categories, and more frequent television exposure, while secondary and tertiary education were negatively associated. Notably, respondents reporting deterioration in past or expected well-being also reported higher verbal satisfaction, suggesting the influence of adaptive reference points or normative response tendencies inherent to verbal satisfaction measures.

In 2021, a markedly different pattern emerged. Female gender and tertiary education showed strong positive associations with Cantril Ladder scores, whereas formerly married status and perceptions of worsened past well-being were strongly negative. Expected future well-being remained consistently associated with evaluative judgments across both waves, although with greater magnitude in 2021.

Taken together, these cross-sectional patterns suggest that evaluative well-being in 2021—measured using the Cantril Ladder—was more strongly differentiated by social position and perceived life trajectory than in 2016. This likely reflects both heightened sensitivity to structural constraints during a period of economic and social strain and the ladder’s greater emphasis on relative social comparison.^23,24^ The consistent importance of future expectations across both years reinforces the role of anticipated life progress as a key driver of evaluative SWB in this age group.^23,25^

At the contextual level, the temporal decline in affective happiness remained robust across models adjusting for institutional quality (Cleanliness Index), state fragility (GVI), and inequality (Gini coefficient), indicating that these macro-level dimensions did not fully account for the post-2016 reduction in youth happiness. While higher state-level inequality (Gini coefficient β = 1.0) and better institutional quality (β = 0.01) were positively associated with happiness, these effects should be interpreted cautiously, particularly given their ecological nature and, in the case of institutional quality, small magnitude. The GVI showed a statistically significant but substantively negligible association. The positive inequality–SWB gradient is consistent with mixed findings in low- and middle-income settings, where inequality does not always reduce SWB and may even correlate positively in contexts of adaptation or strong social capital.^5,26^ This echoes the Easterlin paradox, which posits that within countries wealthier people are happier than poorer ones, but across countries or over time, average happiness does not reliably rise with income or inequality, as relative comparisons and non-material factors dominate.^27–29^

Positive associations between inequality and subjective well-being have been reported in some LMIC and sub-Saharan African contexts and may reflect context-specific mechanisms such as adaptation to inequality, selective migration, remittance flows, or social cohesion within unequal but tightly knit communities, rather than protective effects of inequality per se.^5,26^ The low between-state variation in the mixed-effects robustness check (ICC ≈ 3.4%) further suggests that structural factors operate primarily through individual-level pathways rather than strong clustering at the state level.

Across the contextual models, individual-level predictors were stable and theoretically consistent. Female gender (β ≈ 0.15–0.21) and higher educational attainment were positively associated with affective happiness, consistent with evidence from Nigeria and Kenya highlighting gender and schooling as sources of resilience among youth despite structural disadvantages.^5,23^ Formerly and never-married status were strongly negatively associated with happiness, likely reflecting heightened social and economic vulnerability within this age group. Less frequent television viewing was negatively associated with happiness in some model specifications, potentially capturing reduced access to information, social connectedness, or shared social narratives rather than social comparison per se. Perceptions of worsened past and future well-being emerged as the strongest negative correlates, underscoring the central role of subjective appraisals of life trajectory in shaping youth subjective well-being.^29^

Happiness surveys, as a proxy for population mental health, face methodological challenges such as order bias, mood sensitivity, personality traits, and correlated errors.^30^ While the economics of happiness framework provides valuable insights into structural determinants of well-being^29^ this study positions SWB primarily as a proxy for population mental health rather than purely economic utility. In line with Maslow’s hierarchy of needs — where basic security and belonging precede self-actualization — the strong negative associations with worsened perceived trajectories and formerly married status suggest that unmet lower-level needs (safety, social connection) may underlie much of the observed decline — a framing more aligned with mental health than narrow economic welfare. However, the large sample, consistent happiness item, and survey-weighted estimation in this study help mitigate these concerns, supporting SWB as a feasible and policy-relevant indicator for monitoring youth mental health in resource-constrained settings.^11^

## Strengths, Limitations, and Implications

This study offers several key strengths. First, it draws on nationally representative MICS data from two waves (2016–2017 and 2021), yielding a large, design-weighted sample (n ≈ 41,160) that supports generalizable inferences about youth well-being in Nigeria. Second, the consistent measurement of affective well-being (happiness item identical across waves) enables reliable temporal comparison, while the separate handling of evaluative well-being (due to the 2016 verbal vs 2021 Cantril Ladder change) ensures methodological transparency and avoids invalid pooling. Third, the inclusion of multiple state-level contextual indicators (institutional quality, fragility, inequality) and a Year × GVI interaction provides a nuanced examination of how structural conditions moderate temporal trends — a dimension that remains underexplored in existing Nigerian and SSA youth SWB research. Fourth, the robustness check with a random-intercept mixed-effects model (low ICC ≈ 3.4%) confirms that findings are not driven by state-level clustering, reinforcing the primacy of survey-weighted models.

Despite these strengths, several limitations must be acknowledged. The repeated cross-sectional design precludes estimation of within-person change over time; observed differences may partly reflect shifts in sample composition, cohort effects, or period influences rather than individual trajectories. The measurement change in evaluative well-being prevented direct temporal comparison of life satisfaction, limiting insights into that dimension. Residual confounding from unmeasured variables (e.g., direct exposure to violence, peer support, household shocks, or individual personality traits) remains possible. The unexpected positive ecological association between state-level inequality (Gini) and happiness, while plausible in some LMIC/SSA contexts, warrants cautious interpretation and further replication. Finally, happiness surveys face inherent challenges (order bias, mood sensitivity, personality traits, correlated errors; Bertrand & Mullainathan, 2001; Frey & Stutzer, 2002), though the large sample, consistent item, and survey-weighted estimation help mitigate these concerns.

Despite these limitations, the findings carry important implications for policy and future research. The robust temporal decline in affective happiness, independent of major contextual factors, highlights the the urgency of multisectoral interventions addressing both individual resilience (education, social support, perceived security) and place-based vulnerabilities (governance quality, inequality reduction, fragility mitigation). The amplified negative role of state-level vulnerability in 2021 underscores the need for subnational targeting in high-GVI areas. Future research should prioritize longitudinal/panel designs to capture within-person trajectories, incorporate additional psychosocial mediators (e.g., direct trauma exposure, peer networks), and explore the mechanisms behind the positive Gini association using mixed-methods approaches.

## CONCLUSION

The period 2016–2021 was marked by a clear and persistent decline in affective happiness among Nigerian youth, which was not fully explained by institutional, fragility, or inequality contexts and appeared amplified in more vulnerable states. Research Findings suggestt targeted, evidence-based interventions are urgently needed to reverse this trend and protect the well-being of the next generation in fragile and unequal settings.

## Data Availability Statement

The underlying primary data for this study are derived from harmonized datasets from **IPUMS MICS** (Multiple Indicator Cluster Surveys, Nigeria waves 2016 and 2021) and the Global Data Lab (GDL) state-level vulnerability index. These data are freely available upon registration and request for customized extracts from the IPUMS MICS website and Global Data Lab. Researchers must follow the respective citation guidelines provided by the sources.

The processed dataset (4youth_happiness.rds), R analysis scripts (for data cleaning, descriptive statistics, survey-weighted modeling, robustness checks, and figure generation), and associated outputs (tables, figures, model summaries) are openly available in this Zenodo archive: https://doi.org/10.5281/zenodo.18314154 (version 1.0, published January 20, 2026). This archive corresponds to the GitHub repository at https://github.com/olasuraheem/IPUM_SWB (release v1.0). The materials are provided under the MIT License and include reproduction instructions in the README.md file (requires R ≥ 4.4.1 and listed packages).

## Consent for publication

Not applicable.

## Author Contributions

Kamaldeen Sunkanmi Abdulraheem (KSA) conceived the study, designed the methodology, acquired and processed the data from IPUMS MICS and Global Data Lab, performed all statistical analyses and visualizations, interpreted the results, drafted the manuscript, revised it critically, and approved the final version for submission. The author is responsible for the accuracy and integrity of the work.

## Data Availability

The underlying primary data for this study are derived from harmonized datasets from IPUMS MICS (Multiple Indicator Cluster Surveys, Nigeria waves 2016 and 2021) and the Global Data Lab (GDL) state-level vulnerability index. These data are freely available upon registration and request for customized extracts from the IPUMS MICS website and Global Data Lab. Researchers must follow the respective citation guidelines provided by the sources.
The processed dataset (4youth_happiness.rds), R analysis scripts (for data cleaning, descriptive statistics, survey-weighted modeling, robustness checks, and figure generation), and associated outputs (tables, figures, model summaries) are openly available in this Zenodo archive: https://doi.org/10.5281/zenodo.18314154 (version 1.0, published January 20, 2026). This archive corresponds to the GitHub repository at https://github.com/olasuraheem/IPUM_SWB (release v1.0). The materials are provided under the MIT License and include reproduction instructions in the README.md file (requires R ≥ 4.4.1 and listed packages).

https://doi.org/10.5281/zenodo.18314154

